# Exploring Youth-Perceived Barriers and Attitudes Towards the Clinical Use of Pharmacogenetic Testing to Optimise Antidepressant Pharmacotherapy

**DOI:** 10.1101/2024.08.25.24312539

**Authors:** Bradley Roberts, Zahra Cooper, Georgia Landery, Susanne Stanley, Bernadette T. Majda, Khan, R. L. Collins, P. Anthony Akkari, Sean D. Hood, Jennifer Rodger

## Abstract

The field of pharmacogenetics (PGx) is experiencing significant growth, with increasing evidence to support its application in psychiatric care, suggesting its potential to personalise treatment plans, optimise medication efficacy, and reduce adverse drug reactions. However, the perceived utility and practicability of PGx for psychiatric treatment in youth remains underexplored. This study investigated youth-perceived barriers and attitudes towards the implementation of PGx testing to guide antidepressant treatment in primary care. Semi-structured focus groups and interviews were conducted with 17 participants aged between 18 to 24 years. These sessions were recorded and transcribed before thematic analysis was used to identify collective themes. Three key themes were identified, including attitudes towards the medication prescription process, concerns and attitudes towards PGx testing, and perceived barriers to its clinical implementation. Although PGx testing was positively perceived by most participants, all participants shared concerns about PGx testing. Participants voiced concerns about the financial impact of PGx testing, the potential for treatment delays, and the accuracy of PGx testing in guiding antidepressant treatment. Additionally, participants noted that the low awareness and willingness of general practitioners to incorporate PGx testing into routine practice could hinder successful clinical implementation. Prior to the implementation of PGx testing into Australian primary practices, it is essential to acknowledge patient perspectives and ensure that clinical practices remain patient-focused. This study highlights important considerations for integrating PGx testing into antidepressant pharmacotherapy and emphasizes the need for future research to address and mitigate youth-perceived barriers.

## Introduction

Over one in five Australian young adults under the age of 25 experience symptoms of depression and/or anxiety [1, 2]. Whilst antidepressants are often considered the first-line pharmacotherapy for these conditions [3, 4], these medications have consistently produced poor results in youth patients [5-15], with over one-third of youth treated with antidepressant pharmacotherapy showing little to no response to their antidepressant treatment [16]. Where patients fail to respond to an adequate antidepressant trial, another is often prescribed in a trial-and-error manner [17]. However, the likelihood of future response and remission with antidepressant pharmacotherapy decreases dramatically with subsequent antidepressant trials [18, 19]. Similarly, the risk of severe medication-related side effects, which may significantly impact quality of life, increases with each consecutively prescribed antidepressant. It is therefore crucial to ensure that antidepressants are prescribed optimally from the initial treatment.

The Australian prescription rate of antidepressants has risen by ∼25% per annum over the past 7 years [20-25], with over 33 million antidepressants prescribed in 2023 alone [20]. In recent years, antidepressant use in young Australians under 25 years increased by 25% and new antidepressant prescriptions to those not previously prescribed, rose by over 33% [26]. Given that over 85% of antidepressants in Australia are prescribed by general practitioners (GPs) [20], there is a considerable need to reanalyse primary mental health care practices to improve the effectiveness of antidepressant pharmacotherapy and ensure that youth receive optimal care [27].

Pharmacogenetics (PGx), the study of how interindividual genetic variation affects a person’s response to medication, has been suggested to aid clinicians in the medication selection process, personalising a patients prescription portfolio [28, 29]. Numerous studies have shown that PGx-guided pharmacotherapy improves the therapeutic effect of antidepressant treatment, decreases the likelihood of adverse drug reactions and medication-related side effects, and increases patient medication adherence [30-46]. PGx has also been suggested to have particular relevance in the treatment of depression and anxiety in young people due to the heightened risk profile and low medication adherence rates observed in youth compared to adults [47]. Though support for PGx is still emerging [43, 48-52], the rapidly expanding capabilities for PGx-guided therapeutics could potentially address these challenges, facilitating wider implementation for young people with depression and anxiety [53].

To date, the uptake of PGx-guided pharmacotherapy in practice in Australia has been slow, with previous studies bringing to light several barriers impeding clinical integration [54-56]. Previous research aimed to understand the perspectives of key stakeholders, investigating attitudes towards PGx-guided mental health care among various groups, such as adult patients [57-60], practitioners and healthcare specialists [60-70], and even youth psychiatrists [71-73]. However, little research has evaluated the perspectives of younger patient populations, and those that have, did so only after PGx testing had taken place [74].

Young people are increasingly seeking more involvement in their healthcare decision making [75, 76]. Therefore, it is important to understand their perspectives and views on clinical interventions such as PGx to ensure that their needs and expectations are met during its implementation into clinical use [77]. Given the limited research on youth perspectives, this study aimed to understand young people’s concerns surrounding antidepressant treatment, their attitudes and perspectives on PGx testing in antidepressant pharmacotherapy, and the perceived barriers that may inhibit Australian youth from accessing PGx testing in primary mental healthcare.

## Materials and Methods

### Study Aim and Design

A qualitative approach utilising focus groups and interviews was employed to gain a deeper understanding of young adults’ perspectives of PGx-guided antidepressant pharmacotherapy. This methodology was selected to gather rich, detailed data, providing valuable insights into the nuanced and complex views of participants, often challenging to quantify. Both inductive and deductive coding was used to explore organically emerging themes and those guided by our principal framework.

This study is part of a larger co-design research project consisting of two consecutive stages: (1) a) exploratory focus groups and interviews with young adults with a lived experience of depression and/or anxiety, and b) case study discussions with GPs with an interest in treating youth mental health, and (2) a pilot randomised controlled trial evaluating the effectiveness of PGx-guided antidepressant pharmacotherapy in young people. This iterative design ensures the findings of the focus groups and interviews inform the development of the outcome measures for the pilot randomised controlled trial. This paper will report on the findings obtained from the focus groups and interviews with young adults.

### Participant Recruitment

Purposive sampling was used to recruit participants for focus group and interview discussions. Eligibility criteria included (1) age between the ages of 18 and 24 years inclusive, (2) self-reported current or past antidepressant pharmacotherapy, and (3) fluency in written and spoken English sufficient to provide informed consent. Individuals who were unable to give informed consent due to cognitive or linguistic reasons were excluded from participation. Recruitment methods included advertisements on social media, fliers on university campuses, and snowballing sampling through word-of-mouth.

This study was approved by the Government of Western Australia, Department of Health (RGS0000005473) and endorsed by the University of Western Australia (2023/ET000209). All eligible participants expressing interest were emailed study information letters. Informed consent was obtained from all participants prior to taking part in a focus group or interview discussion along with completion of a participant demographic form. Reminder emails were sent one week prior to the session and repeated 24 hours before the scheduled focus group or interview session. All sessions were held in a private meeting room at the Perron Institute for Neurological and Translational Science, Perth.

### Focus Group and Interview Discussions

Semi-structured focus groups and interviews were conducted to explore the attitudes and perceptions of young people toward PGx testing in antidepressant pharmacotherapy. Where feasible, participant scheduling prioritised focus groups of 2-4 participants to facilitate interactive discussions, allowing for a broader range of perspectives. Where focus groups were not possible due to participant preference or availability, individual interviews were conducted. All focus groups and interviews were moderated by ZC and BR and lasted approximately 60 minutes.

Facilitators followed the principal guide sequentially but allowed flexibility for participants to lead the conversation and contribute to each question. Facilitated prompts were used when discussion waned. Questions in the principal guide were constructed from prior literary review and the aid of community members from the Western Australian Health Translation Network’s Consumer and Community Involvement Program to ensure that the questions asked were targeted and framed appropriately for understanding by the youth demographic [53]. Each session included the eight questions presented in Table 1.

**Table 1.** Principal questions for focus groups and interviews.

|  |
| --- |
| 1. What is your experience in getting treatment for depression and/or anxiety? |
| 2. What are your questions or concerns when starting a new medicine? |
| 3. What do you think affects a person's response to new medication? |
| 4. What do you see as advantages of PGx testing in depression and/or anxiety? |
| 5. What concerns would you have if you were offered DNA testing to select the right medication for your depression or anxiety? |
| 6. Do you foresee any barriers to getting PGx testing for the treatment of depression and/or anxiety? |
| 7. In general, do you find that the information that you received from healthcare professionals (such as GPs) about your anxiety and/or depression and/or medication is adequate? |
| 8. How can GPs offer PGx testing in the treatment of mental health conditions? How can GPs improve the information you receive about your treatment plan? |

### Data Collection

All focus groups and interviews were audio recorded and transcribed using an orthographic style to maintain the accuracy and integrity of the data. Transcripts were edited to preserve the anonymity of participants, removing names, and replacing them with unique identifiers (i.e., FG1P1).

### Data Analysis

Ritchie and Spencer’s Thematic Analysis Framework was used to analyse each focus group and interview session [78]. Given the semi-structured nature of the sessions allowing for participant-led discussions, novel themes were also constructed using reflexive thematic analysis techniques as outlined by Braun and Clarke [79].

First-pass coding was conducted by BR using MS Excel to identify broad-scope ideas expressed by participants in each focus group or interview. This process helped determine whether thematic data saturation had been reached. Second-pass coding grouped these codes into novel or predetermined themes. The data was then independently coded by GL. Differing codes were moderated and finalised by ZC.

## Results

### Participant Demographics

Five focus groups and four interviews were conducted. A total of 32 young people expressed interest and accepted the email invitation to partake in a focus group or interview discussion. Of these, 17 participants met eligibility requirements and attended and participated in either a focus group or interview discussion. Most participants were female (*n* = 12, 70.6%) with a post-high school education (*n* = 12, 70.6%); approximately 52.3% (*n* = 9) of the sample were born outside of Australia. Full demographic details of all participants are provided in Supplementary Table 1.

### Findings

Themes from the focus groups and interviews included statements related to youth experiences and perceptions on current antidepressant prescription procedures and their thoughts and concerns regarding the implementation of PGx testing to inform antidepressant pharmacotherapy. This paper reports on arising themes specifically related to: (1) concerns when starting antidepressant pharmacotherapy, (2) perspectives and concerns surrounding pharmacogenetic testing, and (3) youth-perceived barriers to pharmacogenetic implementation, as these themes relate to the implementation of PGx into clinical practice for youth depression. Key findings are summarised in Supplementary Table 2.

### Concerns When Starting Antidepressant Pharmacotherapy

Youth participants were prompted to share their experiences when receiving antidepressant medication, with all participants sharing at least one major concern they had during this treatment process. Two commonly recuring subthemes arose in all focus groups and interview sessions: 1) lack of involvement in the treatment process, and 2) effectiveness and tolerability.

#### Lack of Involvement in the Treatment Process

Many participants reported a significant concern regarding their involvement in the medication selection process. Specifically, they felt excluded from treatment decisions leading to feelings of disempowerment and frustration, and ultimately, losing trust in their clinician’s treatment decisions.

> *“I told you my preference and you just completely disregarded it. So, I got put on Prozac and my health took a plunge. Like a really, really bad plunge.”*

Furthermore, participants frequently highlighted the impersonal nature of the prescription process, sharing that treatments were often prescribed based on general guidelines rather than investigating individualised needs.

> *“…the way that they prescribe, they had a flip book of medications, and they were like, ‘yeah, do you want that one?’ Like, do I look like I’m here to shop? I’m here to talk about my health.”*

Participants shared the desire to be included in the decision-making process, with strong preferences for a more collaborative approach with healthcare providers and conversations to determine shared expectations and treatment goals. Participants felt that these crucial conversations were often neglected, leaving them unprepared and ill-informed when it came to their antidepressant prescription and mental health treatment.

> *“Like, they don’t really talk to you that much about your background it’s just like, okay, what medication can we try next. So that wasn’t working, let’s try another one. And that’s mainly what my experience consisted of.”*

Participants also discussed their eagerness to engage in conversations regarding their treatment options, including alternative medications and non-pharmacological interventions, allowing them to be better informed and have agency in their treatment.

> *“Whenever I go to the GP, they’re like, ‘oh yeah. Just have your medication. Okay. Goodbye’. I didn’t know any of the other options. So, I would have liked to know, like choices or other things that I could have had.”*

#### Treatment Effectiveness and Tolerability

Almost all participants reported having experienced some form of medication-related side effects, with many participants anxious about future occurrences and the long-term consequences of antidepressants.

> *“…those 2-3 weeks post having started medication was the worst two or three weeks I’ve ever experienced. I was shaking the whole time, didn’t sleep for like weeks. It got worse before it got better in terms of the depression symptoms.”*

Coupled with these experiences, numerous participants also experienced being on antidepressants that had not provided any therapeutic benefit, creating an apprehension for future prescriptions and an expectation that these future treatments would also fail. This uncertainty contributed to the hesitation in starting or continuing with antidepressant pharmacotherapy, with over half the participants from the focus group and interview sessions having not responded to at least one antidepressant medication.

Due to these experiences, participants frequently highlighted the challenge of finding an effective medication, emphasising the exhaustive nature of the trial-and-error process often required to achieve a therapeutic response. Frustrations with the time-consuming and sometimes discouraging process of trying multiple medications along with the uncertainty about whether any given medication would be effective, often led to participants to cease their treatment without telling their treating clinician.

> *“I was on and off medication for four years. I kind of just stopped it because I didn’t see any effects and it just seemed like the GP was just too busy and just disinterested because you only get your 10 minutes. So, I kind of just stopped that by myself.”*

Lastly, several participants feared becoming reliant on their medication to maintain their mental health. This fear was compounded by worries about changes in behavioural traits and personality, with participants questioning whether the medication would alter their identity, dampening aspects of their personality or changing their behaviour in undesirable ways.

> *“…but then I feel like now I’m like starting to want to get off antidepressants because I feel like I don’t even remember what I was like before I went on antidepressants and like, I feel like I’ve changed a lot since then.”*

### Perspectives and Concerns Surrounding Pharmacogenetic Testing

Participants generally viewed PGx testing as a beneficial tool for informing antidepressant prescriptions, recognising that genetic variability can impact drug metabolism and response. The general consensus was that PGx information could be used to optimise and personalise antidepressant prescriptions.

> *“If [PGx testing] was available to me, I would probably go to that in the first instance, just because then I know that it’s, at least somewhat customised to my body and I know that it’s probably going to work. I found it quite a traumatic process of finding which medication works. And I know this would save time, a lot of money, a lot of stress and then not going through those side effects. And also, if you’re able to change the dose to however your body processes medication, you kind of already know where you should be, and it’s only like little minor tweaks that you probably do to the dose as opposed to changing like a whole medication or adding new medications.”*

Despite this positivity, participants shared several concerns they would wish to see addressed prior to PGx use.

#### Financial Impact

Concerns predominantly centred around the costs associated with PGx testing, with participants often perceiving PGx as a financial burden involving frequent visits to the GP to engage with testing processes.

> *“I’m sure genetic testing is very difficult, and this is not a test that I think they can easily just put into a machine, and it spits it out, so I’m presuming it would be quite an expensive test.”*

Several participants grossly overestimated the expense of PGx testing, believing it to be prohibitively high, whereas those who understood the costs involved, recognised that $200 would be a reasonable upper limit for their use.

> *“If I thought it was going to really do well for me, I’d probably be okay with maybe $200. Yeah, but like anything more than that and I would ask, can I afford this.”*

For a few participants, financial anxieties were compounded by familial concerns, with participants worrying that, as a young person reliant on the financial support of family members, the cost of PGx testing would not be favoured by parents when weighed against other medical expenditures.

#### Treatment Delay

Mixed views were shared regarding the potential for PGx testing processes to delay treatment. Whilst some participants perceived any wait time for test results as acceptable if it led to more effective treatment, others were concerned that the delay between sample collection and receiving results would postpone the start of their treatment.

> *“If you need something right away and want to start taking medication, it might take longer for all the things in this process; for them to do the swab, then to send it out, then to have them analyse it and then give the results back and then get the results to you, it might take a while longer.”*
>
> *“I think that the time for me, even if it took like a while, I think it would be worth it to have a more targeted treatment.”*

Commonly, participants concerned about treatment delays due to PGx testing often overestimated the duration of the testing processes, with some expecting results and medication initiation to take over a month. When focus group and interview moderators explained that the average wait time for testing in Australia was 10 business days [80], these concerns were often alleviated, though some participants stated that more than one week is an unacceptable time to wait for PGx results and treatment.

> *“If you could be prescribed something a little bit less for a little bit of relief whilst waiting for testing results, you could probably go like several weeks, maybe up to a month. But if you’re on absolutely nothing, probably not more than a week.”*

#### Perspectives on the Terminology “Pharmacogenetics”

Participants expressed varying views on the term “pharmacogenetics”. Some participants found the term to be “*routine*” and “*appropriate*”, whilst others found it “*scary*”, “*high tech*” and “*confusing*”, saying that they would “*probably just ignore it*” should they have come across it during their treatment journey. For some, the inclusion of the term “*pharma*” and “*genetics*” had a poor connotation, linking PGx with “Big Pharma”.

> *“The term itself brings concerns, because anything that you hear related to genetics, it feels really ‘*sciencey’ *and kind of like you think of like some lab somewhere? It makes me a little bit uncomfortable.”*

Many participants felt that the acronym "PGx" would be better suited for engaging and promoting the use of PGx testing with young people, offering a simpler and more approachable way to refer to the concept. Others thought that the name should be shared with descriptors more commonly used colloquially that may decrease the confusion behind the terminology, such as “*DNA testing*” and “*drug metabolism*”.

#### Testing Accuracy

Having experienced failures with prior medications, many participants were concerned about the ability of PGx testing to effectively inform on a medication that would be more appropriate for them and that may more effectively manage their depressive and/or anxiety symptoms.

> *“I am a bit dubious about the results. Like it seems so left of field that somebody can take a swab from my cheek, and it will tell me that they know the answers to my, you know, my biggest fears.”*

#### Invasiveness

Prior to further understanding of what was involved in the testing process, participants believed that PGx testing was a demanding process for mental health patients.

> *“I think personally, anything related to hospitals and just going in to test and get blood tests, it’s really yeah, the whole thing is scary for me, personally.”*

Data-wise, participants voiced their worries about the security and confidentiality of their genetic information. There was a pervasive anxiety that sensitive genetic data could be misused or accessed by unauthorized parties, leading to potential discrimination in future employment or insurance.

Youth-Perceived Barriers to Pharmacogenetic Testing Implementation

Participants considered the concerns of the broader youth population and suggested factors likely inhibiting the broader use of PGx testing in primary mental health care.

#### Knowledge and Willingness of GPs to Adopt PGx Testing

Participants perceived that the understanding and willingness for GPs to integrate PGx testing would stand as a significant barrier to community access. To youth, the perceived lack of understanding meant that GPs would be unfamiliar with how to prescribe medication from patient genotypes and metabolic phenotypes, limiting the value of PGx in primary practice. Furthermore, many participants felt that GPs were not familiar with or adequately informed about how PGx testing could be used in personalised pharmacotherapies and would therefore choose against adopting PGx processes into their routine practice.

> *“I think even some professionals may not believe in the science of it. I had the test done and took it to my doctor and she was like, ‘oh well, this doesn’t really mean anything.’ So yeah, I don’t know whether there’s some professionals that don’t really believe in its validity still.”*
>
> *“Um doctors, you know, doctors don’t want to try and adopt new things. They want to just like, quickly try and find, like the band aid solution. Not really dig deeper into it.”*

A consensus amongst participants was that if PGx testing was to succeed in integrating into the mental healthcare environment, it would be crucial that GPs are not only aware of PGx testing as an option for informed prescriptive practices, but that GPs also endorse and adopt PGx testing as a practice in their day-to-day clinical care regime.

#### Affordability

For young people, the financial impact of PGx testing is a critical hurdle to overcome. Not only do costs impact upon individual participants and their personal concerns with PGx testing, but the financial burden accompanied with the integration of PGx testing into clinical use was perceived as a significant barrier to widespread access amongst youth. Furthermore, participants assumed that PGx testing would be subsidised by Medicare, waiving or reducing the financial burden of testing. When explained by moderators that Medicare currently does not cover PGx testing, participants expressed adamant concerns that PGx testing would be unaffordable for the youth population, stating that PGx testing “*should not be more than $100*”.

Participants emphasised the need for Medicare coverage to include PGx testing to make it a viable option for more young individuals, particularly those from lower-income backgrounds and for those who may not have the support of families to assist them financially.

> *“Um, and obviously any cost is definitely a barrier. As a young person, cost is the biggest issue.”*

#### Lack of Awareness

Participants noted the lack of awareness and community education around PGx in youth mental health care. Only one participant had previously received a PGx test for guided antidepressant treatment, with one other aware of PGx testing, though they had not pursued it. All other participants admitted to a lack of knowledge about PGx testing and its potential benefits for personalised antidepressant treatment.

> *“Looking back on it now, I wish I’d had PGx testing as an option so that it could help, because your medication choice, it is going to be with you for a very long time.”*

Additionally, there was a perceived absence of comprehensive education initiatives within communities to promote an understanding of genetics and its relevance to mental health care. Young people believed that by focusing attention on overcoming other barriers, such as GP knowledge and adoption, that youth and broader community awareness would follow suit. One participant even speculated that the addition of PGx testing to the Medicare Benefit Schedule could not only prove beneficial for young people financially, but also increase the awareness of PGx testing availability amongst the community.

> *“I feel like if it gets funded by Medicare, it might, probably become common knowledge through, like, media resources as well.”*

#### Limited Accessibility

Many participants thought that geographic disparities in healthcare access could present as a significant barrier to widespread adoption of PGx testing in Western Australia, particularly in rural and remote regions. Participants noted that limited access to testing kits and information about PGx benefits in mental health treatment could exacerbate healthcare inequalities, potentially restricting rural youth from accessing personalised PGx-based therapies.

> *“Especially with rural, that might be a bit of a challenge too, not only access to, you know healthcare clinicians, but to see where this test is available in your area.”*

## Discussion

Incorporating PGx-guided antidepressant treatment may address the concerns of young Australians when starting pharmacotherapy for depression and/or anxiety. Yet, integrating PGx into primary mental health care to guide antidepressant pharmacotherapy does not come without its own concerns and more widespread and prevalent barriers to navigate to ensure successful integration. The focus groups and interviews employed in the present study shone some light on young adults’ PGx-related concerns, highlighting the need to address the financial impact, testing accuracy, and practicality of PGx-guided antidepressant treatment. Furthermore, raising community knowledge and awareness of PGx testing, alleviating the financial burden, and improving testing access, is perceived by young adults to improve the effectiveness of PGx integration in youth mental health care.

Young adults participating in this study expressed their dissatisfaction with their lack of involvement in the decision-making process for their antidepressant treatment. Previous research has found that young people frequently express a strong desire for autonomy, taking higher levels of responsibilities for their health by being better informed of their options and thereby more involved in the decision-making process [75, 81, 82]. The conversations that took place in the present study demonstrate that these needs are not currently being met in primary mental health care practice, with a significant disparity between expected and actual involvement in the treatment decision-making process. While adolescents may benefit from professional support and guidance during medical decision-making processes [83], study participants primarily expressed a desire for more personalised treatment to fit their unique circumstances. The lack of comprehensive discussions in primary care practice shown in this study echoes the call for improved communication between healthcare providers and youth patients [84]. Moreover, prioritising prescribing practices that consider individualised factors for each patient need to be explored to empower youth and satisfy their need for personalised care.

Despite contested opinion as to the use of PGx testing in youth mental health practice, prior studies have shown a general optimism amongst young people towards PGx practices, with adolescents experiencing reassurance from the knowledge gained through the PGx results, appreciating the personalised nature of their treatment [74]. However, little is known about the unique perspectives and concerns of young patients naïve to the practice of PGx testing. The present study showed an overwhelming positivity towards PGx testing for mental health by young adults. However, this positivity was coupled with apprehensions around the perceived high costs, delays in obtaining results and accessing treatment, and privacy issues related to genetic data handling, reflecting broader uncertainties in the field [85, 86]. Participants expressed their concerns that costs exceeding $100 (AUD) and wait times of more than one week would be too much to ask of young people who need immediate treatment for depression or anxiety. Given that current PGx testing processes cost on average $180 (AUD) and take approximately 10 business days [80], improvements to these services will need to be made to increase the likelihood of youth engagement in PGx.

Though recent trials have aimed to determine the efficacy and tolerability of PGx-guided antidepressant treatment in young people, the effectiveness of such treatment remains nuanced and further work is required to understand the implications of PGx testing in youth mental health before their concerns can be [43, 51, 52]. Furthermore, future trials must look beyond measures of efficacy and tolerability, assessing patient satisfaction, cost-effectiveness, and treatment adherence in relation to PGx-guided treatment antidepressant treatment, allowing clinicians and researchers alike to gain a broader perspective of the holistic impact of PGx testing. Understanding the concerns of youth is an initial step and offers a unique platform for future research, identifying key outcome measures for future studies, ensuring that trial procedures and findings answer the questions and queries raised by young people.

Youth-perceived barriers identified in this study include a perceived lack of awareness and education about PGx testing among both healthcare providers and the public, financial constraints due to high testing costs, and social and geographical limitations to testing access. These youth-perceived barriers align with those expressed by adults in the literature [54-56]. Youth being unaware of PGx testing as an available option for informed prescription presents as a significant inhibitor to successfully integrating PGx testing into clinical care and vital efforts need to be made to increase this community understanding. Additionally, these efforts are needed to be made to educate and inform regardless of geographical location. Australia is a geographically large nation with over 30% of the nation’s people living outside of metropolitan areas, and thus, PGx needs to be made equitably accessible to all those living in rural environments. Several PGx testing companies in Australia utilise a mail service model that navigates this issue, however, it is important that randomised controlled trials include a geographically diverse sample population to ensure the benefits of PGx-guided treatment are observed outside of metropolitan regions. As mentioned by a participant of this study, with further trial evidence supporting the efficacy of PGx-guided antidepressant pharmacotherapy and an increased awareness and demand for personalised mental health care, policy reform to subsidise the financial burden of PGx testing may be amenable.

There are some limitations to the present study. Participants were recruited predominantly through advertisement on university campuses; thus, our findings may not be reflective of the concerns and perceived barriers of young adults who have not pursued tertiary education. Furthermore, the gender representation in our sample is female biased and the concerns of males receiving antidepressant treatment may be underrepresented in our findings. This is not surprising, however, considering the low rate of help-seeking in young males suffering mental health conditions and the higher rate of mental illness in adolescent and young adult females [87]. Lastly, this study was conducted in Western Australia, which may have specific healthcare practices and cultural attitudes that differ from other regions, affecting the applicability of the findings to other contexts.

## Conclusion

In conclusion, this study highlights the unique perspectives and concerns of young adults towards the implementation of PGx testing to optimise and guide antidepressant pharmacotherapy. Future studies must continue to employ a holistic approach, prioritising these concerns when considering best practice for the treatment of mental health in young people. Similarly, fundamental research to educate and inform the community on PGx practices needs to be prioritised. By addressing these concerns and barriers in future research, healthcare providers and researchers can work towards a more personalized and effective approach to antidepressant pharmacotherapy, ultimately improving outcomes for young adults struggling with mental health issues.

## Data Availability

All data produced in the present study are available upon reasonable request to the authors on a case-by-case basis.

## References

1. Australian Institute of Health and Welfare. Australia’s youth: Mental illness. 2021 [Available from: https://www.aihw.gov.au/reports/children-youth/mental-illness.

2. Lawrence D, Johnson S, Hafekost J, Boterhoven de Haan K, Sawyer M, Ainley J, et al. The mental health of children and adolescents. Report on the second Australian child and adolescent survey of mental health and wellbeing. Canberra, ACT, Australia 2015 [Available from: https://www1.health.gov.au/internet/main/publishing.nsf/Content/mental-pubsm-child2

3. Malhi GS, Bassett D, Boyce P, Bryant R, Fitzgerald PB, Fritz K, et al. Royal Australian and New Zealand College of Psychiatrists clinical practice guidelines for mood disorders. Aust N Z J Psychiatry. 2015;49(12):1087–206. 10.1177/0004867415617657.

4. Santarsieri D, Schwartz TL. Antidepressant efficacy and side-effect burden: a quick guide for clinicians. Drugs Context. 2015;4:212290. 10.7573/dic.212290.

5. Strawn JR, Mills JA, Suresh V, Mayes T, Gentry MT, Trivedi M, et al. The impact of age on antidepressant response: A mega-analysis of individuals with major depressive disorder. J Psychiatr Res. 2023;159:266–73. 10.1016/j.jpsychires.2023.01.043.

6. Bridge JA, Iyengar S, Salary CB, Barbe RP, Birmaher B, Pincus HA, et al. Clinical response and risk for reported suicidal ideation and suicide attempts in pediatric antidepressant treatment: a meta-analysis of randomized controlled trials. Jama. 2007;297(15):1683–96. 10.1001/jama.297.15.1683.

7. March J, Silva S, Petrycki S, Curry J, Wells K, Fairbank J, et al. Fluoxetine, cognitive-behavioral therapy, and their combination for adolescents with depression: Treatment for Adolescents With Depression Study (TADS) randomized controlled trial. Jama. 2004;292(7):807–20. 10.1001/jama.292.7.807.

8. Weihs KL, Murphy W, Abbas R, Chiles D, England RD, Ramaker S, et al. Desvenlafaxine versus placebo in a fluoxetine-referenced study of children and adolescents with major depressive disorder. J Child Adolesc Psychopharmacol. 2018;28(1):36–46. 10.1089/cap.2017.0100.

9. Emslie GJ, Prakash A, Zhang Q, Pangallo BA, Bangs ME, March JS. A double-blind efficacy and safety study of duloxetine fixed doses in children and adolescents with major depressive disorder. J Child Adolesc Psychopharmacol. 2014;24(4):170–9. 10.1089/cap.2013.0096.

10. Durgam S, Chen C, Migliore R, Prakash C, Edwards J, Findling RL. A phase 3, double-blind, randomized, placebo-controlled study of vilazodone in adolescents with major depressive disorder. Paediatr Drugs. 2018;20(4):353–63. 10.1007/s40272-018-0290-4.

11. Le Noury J, Nardo JM, Healy D, Jureidini J, Raven M, Tufanaru C, et al. Restoring Study 329: efficacy and harms of paroxetine and imipramine in treatment of major depression in adolescence. Bmj. 2015;351:h4320. 10.1136/bmj.h4320.

12. Davey CG, Chanen AM, Hetrick SE, Cotton SM, Ratheesh A, Amminger GP, et al. The addition of fluoxetine to cognitive behavioural therapy for youth depression (YoDA-C): a randomised, double-blind, placebo-controlled, multicentre clinical trial. Lancet Psychiatry. 2019;6(9):735–44. 10.1016/s2215-0366(19)30215-9.

13. Safer DJ, Zito JM. Short- and long-term antidepressant clinical trials for major depressive disorder in youth: findings and concerns. Front Psychiatry. 2019;10:705. 10.3389/fpsyt.2019.00705.

14. Spielmans GI, Gerwig K. The efficacy of antidepressants on overall well-being and self-reported depression symptom severity in youth: a meta-analysis. Psychother Psychosom. 2014;83(3):158–64. 10.1159/000356191.

15. Cipriani A, Zhou X, Del Giovane C, Hetrick SE, Qin B, Whittington C, et al. Comparative efficacy and tolerability of antidepressants for major depressive disorder in children and adolescents: a network meta-analysis. Lancet. 2016;388(10047):881–90. 10.1016/s0140-6736(16)30385-3.

16. Kennard BD, Silva SG, Tonev S, Rohde P, Hughes JL, Vitiello B, et al. Remission and recovery in the Treatment for Adolescents with Depression Study (TADS): acute and long-term outcomes. J Am Acad Child Adolesc Psychiatry. 2009;48(2):186–95. 10.1097/CHI.0b013e31819176f9.

17. Leuchter AF, Cook IA, Hunter AM, Korb AS. A new paradigm for the prediction of antidepressant treatment response. Dialogues Clin Neurosci. 2009;11(4):435–46. 10.31887/DCNS.2009.11.4/afleuchter.

18. Rush AJ, Trivedi MH, Wisniewski SR, Nierenberg AA, Stewart JW, Warden D, et al. Acute and longer-term outcomes in depressed outpatients requiring one or several treatment steps: a STAR*D report. Am J Psychiatry. 2006;163(11):1905–17. 10.1176/ajp.2006.163.11.1905.

19. Gaynes BN, Warden D, Trivedi MH, Wisniewski SR, Fava M, Rush AJ. What did STAR*D teach us? Results from a large-scale, practical, clinical trial for patients with depression. Psychiatr Serv. 2009;60(11):1439–45. 10.1176/ps.2009.60.11.1439.

20. Australian Institute of Health and Welfare. Mental helath-related prescriptions. 2023 [Available from: https://www.aihw.gov.au/mental-health/topic-areas/mental-health-prescriptions.

21. Australian Institute of Health and Welfare. Mental helath-related prescriptions. Mental health 2022.

22. Australian Institute of Health and Welfare. Mental helath-related prescriptions. Mental health 2021.

23. Australian Institute of Health and Welfare. Mental helath-related prescriptions. Mental health 2020.

24. Australian Institute of Health and Welfare. Mental helath-related prescriptions. Mental health 2019.

25. Australian Institute of Health and Welfare. Mental helath-related prescriptions. Mental health 2018.

26. de Oliveira Costa J, Gillies MB, Schaffer AL, Peiris D, Zoega H, Pearson SA. Changes in antidepressant use in Australia: A nationwide analysis (2015-2021). Aust N Z J Psychiatry. 2023;57(1):49–57. 10.1177/00048674221079740.

27. Le LK, Shih S, Richards-Jones S, Chatterton ML, Engel L, Stevenson C, et al. The cost of Medicare-funded medical and pharmaceutical services for mental disorders in children and adolescents in Australia. PLoS One. 2021;16(4). 10.1371/journal.pone.0249902.

28. Bousman CA, Maruf AA, Marques DF, Brown LC, Müller DJ. The emergence, implementation, and future growth of pharmacogenomics in psychiatry: a narrative review. Psychol Med. 2023;53(16):7983–93. 10.1017/s0033291723002817.

29. Strawn JR, Mills JA, Poweleit EA, Ramsey LB, Croarkin PE. Adverse effects of antidepressant medications and their management in children and adolescents. Pharmacotherapy. 2023. 10.1002/phar.2767.

30. Hall-Flavin DK, Winner JG, Allen JD, Jordan JJ, Nesheim RS, Snyder KA, et al. Using a pharmacogenomic algorithm to guide the treatment of depression. Transl Psychiatry. 2012;2(10):e172. 10.1038/tp.2012.99.

31. Hall-Flavin DK, Winner JG, Allen JD, Carhart JM, Proctor B, Snyder KA, et al. Utility of integrated pharmacogenomic testing to support the treatment of major depressive disorder in a psychiatric outpatient setting. Pharmacogenet Genomics. 2013;23(10):535–48. 10.1097/FPC.0b013e3283649b9a.

32. Greden JF, Parikh SV, Rothschild AJ, Thase ME, Dunlop BW, DeBattista C, et al. Impact of pharmacogenomics on clinical outcomes in major depressive disorder in the GUIDED trial: A large, patient- and rater-blinded, randomized, controlled study. J Psychiatr Res. 2019;111:59–67. 10.1016/j.jpsychires.2019.01.003.

33. Oslin DW, Lynch KG, Shih MC, Ingram EP, Wray LO, Chapman SR, et al. Effect of pharmacogenomic testing for drug-gene Interactions on medication selection and remission of symptoms in major depressive disorder: The PRIME care randomized clinical trial. Jama. 2022;328(2):151–61. 10.1001/jama.2022.9805.

34. Han C, Wang SM, Bahk WM, Lee SJ, Patkar AA, Masand PS, et al. A pharmacogenomic-based antidepressant treatment for patients with major depressive disorder: results from an 8-week, randomized, single-blinded clinical trial. Clin Psychopharmacol Neurosci. 2018;16(4):469–80. 10.9758/cpn.2018.16.4.469.

35. Vos CF, ter Hark SE, Schellekens AFA, Spijker J, van der Meij A, Grotenhuis AJ, et al. Effectiveness of genotype-specific tricyclic antidepressant dosing in patients with major depressive disorder: A randomized clinical Trial. JAMA Network Open. 2023;6(5):e2312443-e. 10.1001/jamanetworkopen.2023.12443.

36. Pérez V, Salavert A, Espadaler J, Tuson M, Saiz-Ruiz J, Sáez-Navarro C, et al. Efficacy of prospective pharmacogenetic testing in the treatment of major depressive disorder: results of a randomized, double-blind clinical trial. BMC Psychiatry. 2017;17(1):250. 10.1186/s12888-017-1412-1.

37. Bradley P, Shiekh M, Mehra V, Vrbicky K, Layle S, Olson MC, et al. Improved efficacy with targeted pharmacogenetic-guided treatment of patients with depression and anxiety: A randomized clinical trial demonstrating clinical utility. J Psychiatr Res. 2018;96:100–7. 10.1016/j.jpsychires.2017.09.024.

38. Brown LC, Stanton JD, Bharthi K, Maruf AA, Müller DJ, Bousman CA. Pharmacogenomic testing and depressive symptom remission: A systematic review and meta-analysis of prospective, controlled clinical trials. Clin Pharmacol Ther. 2022;112(6):1303–17. 10.1002/cpt.2748.

39. Bousman CA, Arandjelovic K, Mancuso SG, Eyre HA, Dunlop BW. Pharmacogenetic tests and depressive symptom remission: a meta-analysis of randomized controlled trials. Pharmacogenomics. 2019;20(1):37–47. 10.2217/pgs-2018-0142.

40. Rosenblat JD, Lee Y, McIntyre RS. The effect of pharmacogenomic testing on response and remission rates in the acute treatment of major depressive disorder: A meta-analysis. J Affect Disord. 2018;241:484–91. 10.1016/j.jad.2018.08.056.

41. Wang X, Wang C, Zhang Y, An Z. Effect of pharmacogenomics testing guiding on clinical outcomes in major depressive disorder: a systematic review and meta-analysis of RCT. BMC Psychiatry. 2023;23(334). 10.1186/s12888-023-04756-2.

42. Olson MC, Maciel A, Gariepy JF, Cullors A, Saldivar JS, Taylor D, et al. Clinical impact of pharmacogenetic-guided treatment for patients exhibiting neuropsychiatric disorders: A randomized controlled trial. Prim Care Companion CNS Disord. 2017;19(2). 10.4088/PCC.16m02036.

43. Nooraeen S, Croarkin PE, Geske JR, Shekunov J, Orth SS, Romanowicz M, et al. High probability of gene-drug interactions associated with medication side effects in adolescent depression: Results from a randomized controlled trial of pharmacogenetic testing. J Child Adolesc Psychopharmacol. 2024;34(1):28–33. 10.1089/cap.2023.0043.

44. Skokou M, Karamperis K, Koufaki MI, Tsermpini EE, Pandi MT, Siamoglou S, et al. Clinical implementation of preemptive pharmacogenomics in psychiatry. EBioMedicine. 2024;101:105009. 10.1016/j.ebiom.2024.105009.

45. Platona RI, Voiță-Mekeres F, Tudoran C, Tudoran M, Enătescu VR. The contribution of genetic testing in optimizing therapy for patients with recurrent depressive disorder. Clinics and Practice. 2024;14(3):703–17.

46. Xu L, Li L, Wang Q, Pan B, Zheng L, Lin Z. Effect of pharmacogenomic testing on the clinical treatment of patients with depressive disorder: A randomized clinical trial. J Affect Disord. 2024;359:117–24. 10.1016/j.jad.2024.05.063.

47. Gast A, Mathes T. Medication adherence influencing factors-an (updated) overview of systematic reviews. Syst Rev. 2019;8(1):112. 10.1186/s13643-019-1014-8.

48. Ariefdjohan M, Lee YM, Stutzman DL, LeNoue S, Wamboldt MZ. The utility of pharmacogenetic-guided psychotropic medication selection for pediatric patients: A retrospective study. Pediatr Rep. 2021;13(3):421–33. 10.3390/pediatric13030049.

49. Aldrich SL, Poweleit EA, Prows CA, Martin LJ, Strawn JR, Ramsey LB. Influence of CYP2C19 metabolizer status on escitalopram/citalopram tolerability and response in youth with anxiety and depressive disorders. Front Pharmacol. 2019;10:99. 10.3389/fphar.2019.00099.

50. Poweleit EA, Aldrich SL, Martin LJ, Hahn D, Strawn JR, Ramsey LB. Pharmacogenetics of sertraline tolerability and response in pediatric anxiety and depressive disorders. J Child Adolesc Psychopharmacol. 2019;29(5):348–61. 10.1089/cap.2019.0017.

51. Namerow LB, Ramsey LB, Malik S, Cortese S, Strawn JR. Editorial: Beyond red light, green light: Examining the role of pharmacogenomics in evidence-based care in child and adolescent psychiatry. J Am Acad Child Adolesc Psychiatry. 2022;61(1):29–31. 10.1016/j.jaac.2021.11.001.

52. Vande Voort JL, Orth SS, Shekunov J, Romanowicz M, Geske JR, Ward JA, et al. A randomized controlled trial of combinatorial pharmacogenetics testing in adolescent depression. J Am Acad Child Adolesc Psychiatry. 2022;61(1):46–55. 10.1016/j.jaac.2021.03.011.

53. Roberts B, Cooper Z, Lu S, Stanley S, Majda BT, Collins KRL, et al. Utility of pharmacogenetic testing to optimise antidepressant pharmacotherapy in youth: a narrative literature review. Frontiers in Pharmacology. 2023;14. 10.3389/fphar.2023.1267294.

54. Jameson A, Fylan B, Bristow GC, Sagoo GS, Dalton C, Cardno A, et al. What are the barriers and enablers to the implementation of pharmacogenetic testing in mental health care settings? Front Genet. 2021;12:740216. 10.3389/fgene.2021.740216.

55. Pinzón-Espinosa J, van der Horst M, Zinkstok J, Austin J, Aalfs C, Batalla A, et al. Barriers to genetic testing in clinical psychiatry and ways to overcome them: from clinicians’ attitudes to sociocultural differences between patients across the globe. Transl Psychiatry. 2022;12(1):442. 10.1038/s41398-022-02203-6.

56. Virelli CR, Mohiuddin AG, Kennedy JL. Barriers to clinical adoption of pharmacogenomic testing in psychiatry: a critical analysis. Translational Psychiatry. 2021;11(1):509. 10.1038/s41398-021-01600-7.

57. Kastrinos A, Campbell-Salome G, Shelton S, Peterson EB, Bylund CL. PGx in psychiatry: Patients’ knowledge, interest, and uncertainty management preferences in the context of pharmacogenomic testing. Patient Educ Couns. 2021;104(4):732–8. 10.1016/j.pec.2020.12.021.

58. Liko I, Lai E, Griffin RJ, Aquilante CL, Lee YM. Patients’ perspectives on psychiatric pharmacogenetic testing. Pharmacopsychiatry. 2020;53(6):256–61. 10.1055/a-1183-5029.

59. McCarthy MJ, Chen Y, Demodena A, Leckband SG, Fischer E, Golshan S, et al. A prospective study to determine the clinical utility of pharmacogenetic testing of veterans with treatment-resistant depression. J Psychopharmacol. 2021;35(8):992–1002. 10.1177/02698811211015224.

60. Slomp C, Morris E, Edwards L, Hoens AM, Landry G, Riches L, et al. Pharmacogenomic testing for major depression: A qualitative study of the perceptions of people with lived experience and professional stakeholders. Can J Psychiatry. 2022:7067437221140383. 10.1177/07067437221140383.

61. Chan CY, Chua BY, Subramaniam M, Suen EL, Lee J. Clinicians’ perceptions of pharmacogenomics use in psychiatry. Pharmacogenomics. 2017;18(6):531–8. 10.2217/pgs-2016-0164.

62. Shishko I, Almeida K, Silvia RJ, Tataronis GR. Psychiatric pharmacists’ perception on the use of pharmacogenomic testing in the mental health population. Pharmacogenomics. 2015;16(9):949–58. 10.2217/pgs.15.22.

63. Aboelbaha S, Zolezzi M, Abdallah O, Eltorki Y. Mental health prescribers’ perceptions on the use of pharmacogenetic testing in the management of depression in the middle East and North Africa region. Pharmgenomics Pers Med. 2023;16:503–18. 10.2147/pgpm.S410240.

64. Vest BM, Wray LO, Brady LA, Thase ME, Beehler GP, Chapman SR, et al. Primary care and mental health providers’ perceptions of implementation of pharmacogenetics testing for depression prescribing. BMC Psychiatry. 2020;20(1):518. 10.1186/s12888-020-02919-z.

65. Lanktree MB, Zai G, Vanderbeek LE, Giuffra DE, Smithson DS, Kipp LB, et al. Positive perception of pharmacogenetic testing for psychotropic medications. Hum Psychopharmacol. 2014;29(3):287–91. 10.1002/hup.2383.

66. Thompson C, Steven PH, Catriona H. Psychiatrist attitudes towards pharmacogenetic testing, direct-to-consumer genetic testing, and integrating genetic counseling into psychiatric patient care. Psychiatry Res. 2015;226(1):68–72. 10.1016/j.psychres.2014.11.044.

67. Dunbar L, Butler R, Wheeler A, Pulford J, Miles W, Sheridan J. Clinician experiences of employing the AmpliChip® CYP450 test in routine psychiatric practice. J Psychopharmacol. 2012;26(3):390–7. 10.1177/0269881109106957.

68. Goodspeed A, Kostman N, Kriete TE, Longtine JW, Smith SM, Marshall P, et al. Leveraging the utility of pharmacogenomics in psychiatry through clinical decision support: a focus group study. Ann Gen Psychiatry. 2019;18:13. 10.1186/s12991-019-0237-3.

69. Laplace B, Calvet B, Lacroix A, Mouchabac S, Picard N, Girard M, et al. Acceptability of pharmacogenetic testing among French psychiatrists, a national survey. J Pers Med. 2021;11(6). 10.3390/jpm11060446.

70. Sperber NR, Roberts MC, Gonzales S, Bendz LM, Cragun D, Haga SB, et al. Pharmacogenetic testing in primary care could bolster depression treatment: A value proposition. Clin Transl Sci. 2024;17(6):e13837. 10.1111/cts.13837.

71. Soda T, Merner AR, Small BJ, Torgerson LN, Muñoz K, Austin J, et al. Child and adolescent psychiatrists’ use, attitudes, and understanding of genetic testing and pharmacogenetics in clinical practice. Psychiatry Res. 2023;325:115246. 10.1016/j.psychres.2023.115246.

72. Liko I, Lee YM, Stutzman DL, Blackmer AB, Deininger KM, Reynolds AM, et al. Providers’ perspectives on the clinical utility of pharmacogenomic testing in pediatric patients. Pharmacogenomics. 2021;22(5):263–74. 10.2217/pgs-2020-0112.

73. Jessel CD, Al Maruf A, Oomen A, Arnold PD, Bousman CA. Pharmacogenetic testing knowledge and attitudes among pediatric psychiatrists and pediatricians in Alberta, Canada. J Can Acad Child Adolesc Psychiatry. 2022;31(1):18–27.

74. Stancil SL, Berrios C, Abdel-Rahman S. Adolescent perceptions of pharmacogenetic testing. Pharmacogenomics. 2021;22(6):335–43. 10.2217/pgs-2020-0177

75. Fleary SA, Joseph P. Adolescents’ health literacy and decision-making: A qualitative study. Am J Health Behav. 2020;44(4):392–408. 10.5993/ajhb.44.4.3.

76. Simmons MB, Hetrick SE, Jorm AF. Experiences of treatment decision making for young people diagnosed with depressive disorders: a qualitative study in primary care and specialist mental health settings. BMC Psychiatry. 2011;11:194. 10.1186/1471-244x-11-194.

77. Subasri M, Cressman C, Arje D, Schreyer L, Cooper E, Patel K, et al. Translating precision health for pediatrics: A scoping review. Children (Basel). 2023;10(5). 10.3390/children10050897.

78. Ritchie J, Spencer L. Qualitative data analysis for applied policy research. The Qualitative Researcher’s Companion. 1994:305–29.

79. Braun V, Clarke V. Using thematic analysis in psychology. Qualitative Research in Psychology. 2006;3(2):77–101. 10.1191/1478088706qp063oa.

80. Forbes M, Hopwood M, Bousman CA. CYP2D6 and CYP2C19 variant coverage of commercial antidepressant pharmacogenomic testing panels available in Victoria, Australia. Genes (Basel). 2023;14(10). 10.3390/genes14101945.

81. Freeman JL, Caldwell PHY, Bennett PA, Scott KM. How adolescents search for and appraise online health information: A systematic review. J Pediatr. 2018;195:244–55.e1. 10.1016/j.jpeds.2017.11.031.

82. Freeman JL, Caldwell PHY, Scott KM. The Role of Trust When Adolescents Search for and Appraise Online Health Information. J Pediatr. 2020;221:215–23.e5. 10.1016/j.jpeds.2020.02.074.

83. Grootens-Wiegers P, Hein IM, van den Broek JM, de Vries MC. Medical decision-making in children and adolescents: developmental and neuroscientific aspects. BMC Pediatr. 2017;17(1):120. 10.1186/s12887-017-0869-x.

84. Patak L, Wilson-Stronks A, Costello J, Kleinpell RM, Henneman EA, Person C, et al. Improving patient-provider communication: a call to action. J Nurs Adm. 2009;39(9):372–6. 10.1097/NNA.0b013e3181b414ca.

85. Joly Y, Saulnier KM, Osien G, Knoppers BM. The ethical framing of personalized medicine. Curr Opin Allergy Clin Immunol. 2014;14(5):404–8. 10.1097/aci.0000000000000091.

86. Young C, MacDougall D. CADTH Horizon Scans. An overview of pharmacogenomic testing for psychiatric disorders: CADTH horizon scan. Ottawa (ON): Canadian Agency for Drugs and Technologies in Health; 2023.

87. Australian Bureau of Statistics. National Study of Mental Health and Wellbeing. ABS; 2022 [Available from: https://www.abs.gov.au/statistics/health/mental-health/national-study-mental-health-and-wellbeing/latest-release.

