## Supplementary Material for "Exploring Youth-Perceived Barriers and Attitudes Towards the Clinical Use of Pharmacogenetic Testing to Optimise Antidepressant Pharmacotherapy"

| **Supplementary Table 1. Participant demographic data** | |
| --- | --- |
| **Characteristic** | ***n* (%^†^)** |
| Sex (*n* = 17) |  |
| Female | 12 (71) |
| Male | 4 (23) |
| Non-Binary / Gender Diverse | 1 (6) |
| Level of Education (*n* = 17) |  |
| Completed High School | 5 (29) |
| Completing Undergraduate Degree | 7 (41) |
| Completed Undergraduate Degree | 3 (18) |
| Completing Postgraduate Degree | 2 (12) |
| Place of Birth (*n* = 17) |  |
| Within Australia | 8 (47) |
| Overseas | 9 (53) |
| Both Parents Born Outside Australia (*n* = 16) |  |
| No | 8 (50) |
| Yes | 8 (50) |
| Language Other Than English Spoken at Home (*n* = 17) |  |
| No | 10 (59) |
| Yes | 7 (41) |
| **^†^**Percentages have been rounded to the whole number. Table columns therefore approximate 100%. | |

| **Supplementary Table 2. Summary of the thematic results from youth participants relating to their perspectives on the implementation to PGx integration in primary psychiatric care.** | |
| --- | --- |
| **Theme** | **Findings** |
| **Concerns when starting antidepressant pharmacotherapy** | |
| *Lack of involvement in the treatment process* | - Participant was not involved in the medication selection process - Prescription process was not personalised to the individual and was made based on general medication guidelines - Participant expressed the desire for more personalised an streamlined treatment - Participant expressed the desire to be included in the medication selection process - No discussion about medication and the possible side effects that may present was had between patient and clinician before antidepressants were prescribed - Participant expressed the desire to hear about alternatives to pharmacotherapy before starting their antidepressant prescription |
| *Treatment effectiveness and tolerability* | - Presentation of medication-related side effects - Dependency on antidepressant medication - Efficacy of drug to achieve a therapeutic response - Trial-and-error process of finding the right medication - Effects on behavioural traits and personality |
| **Perspectives and concerns surrounding pharmacogenetic testing** | |
| *Financial impact* | - Cost concern for PGx testing - Overestimation of PGx testing costs - Financial and familial concerns specific to young people - The costs associated with frequent visits to GPs - Appropriate estimation of PGx testing costs |
| *Treatment delay* | - Delay in care with PGx testing - No concern over wait time - Overestimation of PGx testing wait time - Scepticism around the logistics of PGx testing - Over a one-week wait is harmful - Mental healthcare system wait times are long anyway so waiting |
| *Perspectives on the terminology “pharmacogenetics”* | - Appropriate and medical sounding - Scary and confusing - Link to “Big Pharma” - PGx acronym is more appropriate for the general public - Name needs more explanation |
| *Testing accuracy* | - Efficacy of PGx testing |
| *Invasiveness* | - Physical invasiveness of PGx testing - Invasiveness on data privacy |
| **Youth-perceived barriers to pharmacogenetic testing implementation** | |
| *Knowledge and willingness of GPs to adopt PGx testing* | - Lack of GP understanding about PGx testing and its value in clinical practice - GPs may be unwilling to adopt and implement the recommendations provided by PGx testing |
| *Affordability* | - High costs that are currently uninsured by Medicare or private health insurance |
| *Lack of awareness* | - Lack of awareness of PGx in youth - Lack of community education around genetic testing |
| *Limited accessibility* | - Lack of easy access to testing and information on PGx testing - People in rural regions may find it difficult to get access to PGx testing kits |
| *Things that may promote the use and awareness of PGx testing in young people* | - Having support from family would counteract barrier to PGx - Increasing the ease of access to gain a PGx test - Addition to the Medicare Benefit Schedule will increase awareness of PGx testing - Clinical endorsement for all mental healthcare professionals will help to raise awareness of PGx testing - The desire for effective treatment outweighs any potential barriers to PGx testing |
